# Surface Markers on Supermeres Outperform Extracellular Vesicles in Cancer Diagnosis

**DOI:** 10.1101/2025.09.30.25337017

**Authors:** Sonu Kumar, John Alex Sinclair, Tiger Shi, Gaeun Kim, Runyao Zhu, Grace Gasper, Yichun Wang, James N. Higginbotham, Qin Zhang, Dennis K. Jeppesen, Oleg Tutanov, Maxwell Hamilton, Jeffrey L. Franklin, Al Charest, Robert J. Coffey, Satyajyoti Senapati, Hsueh-Chia Chang

**Affiliations:** Department of Chemical and Biomolecular Engineering, University of Notre Dame, Notre Dame, IN 46556, USA; Department of Medicine, Vanderbilt University Medical Center, Nashville, TN 37232, USA; Department of Cell and Developmental Biology, Vanderbilt University School of Medicine, Nashville, TN 37232, USA; Program in Cancer Biology, Vanderbilt University School of Medicine, Nashville, TN, 37232, USA; Department of Medicine, Beth Israel Deaconess Medical Center, Harvard Medical School, Boston, MA, USA; Cancer Research Institute, Beth Israel Deaconess Medical Center, Boston, MA, USA

**Keywords:** Anion-exchange membrane, Biosensor, Colorectal cancer, Electrokinetics, Supermeres

## Abstract

Extracellular nanocarriers, such as extracellular vesicles (EVs), lipoproteins, supermeres, and exomeres are diverse lipid-protein-nucleic acid assemblies. Among them, supermeres hold significant diagnostic potential but are challenging to characterize due to limited surface biomarker information and labor-intensive isolation methods. This study introduces an isolation-free Ion Exchange Membrane Sensing method for detection of supermeres within 30 minutes using 50 μL of sample, with a sensitivity of 10^6^–10^7^ supermeres/mL. Validation through ultracentrifugation and surface plasmon resonance confirms the detection accuracy and specificity. Supermeres carry key proteins such as HSPA13, ENO2, and DDR1 analogous to tetraspanin in EV. Supermeres outperform sEVs and exomeres across multiple shared and unique surface proteins critical to colorectal cancer diagnosis, highlighting their superior clinical utility and potential as next-generation biomarkers in precision medicine.

## 1. Introduction

Extracellular nanocarriers have emerged as pivotal mediators of intercellular communication and promising biomarkers for a variety of diseases, prompting extensive investigations into their diverse subtypes (*1-8*). This heterogeneous family of nanoparticles comprises classical EVs (Extracellular Vesicles)—including exosomes (30–150 nm, of endosomal origin) (*9,10*) and microvesicles (100–1000 nm, derived from the plasma membrane) (*11-14*) —as well as other nanoscale entities present in biofluids. Among these are lipoproteins (e.g., HDL [∼5–12 nm] (*15-17*) and LDL [∼18–28 nm] (*18-20*) and newly identified extracellular nanoparticles that lack a surrounding membrane (*9,21,22*). Notably, exomeres (∼30–50 nm), recently characterized as non-membranous particles with distinct molecular signatures (*9*), and the even smaller supermeres (*21*) (∼15–25 nm), isolated from the supernatant of exomere preparations (*22*), further underscore the complexity of the secretome. These amembranous entities, which do not conform to traditional EV or lipoprotein categories, expand our understanding of cell-secreted particulate diversity (*13,14,23*). Importantly, while classical EVs have been extensively studied (*15,16,24*), supermeres remain underexplored primarily due to the lack of characterization tools and lack of canonical markers, thereby presenting both a challenge and an opportunity for novel biomarker discovery (*21,25-28*).

Supermeres represent a novel class of secreted particles with a unique biochemical composition and potentially significant biological functions (*21,25,26*). Initial studies have demonstrated that supermeres harbor a rich repertoire of proteins and RNAs that are distinct from those found in small EVs or exomeres (*21*). Comprehensive proteomic analyses have revealed an abundance of disease-related biomolecules within supermeres, including shed ectodomains of membrane protein fragments and growth factor regulators (*21*). For instance, transforming growth factor beta induced (TGFBI) emerged as one of the predominant proteins in supermeres isolated from colorectal cancer models, while other oncogenic elements—such as extracellular fragments of transmembrane proteins like the epidermal growth factor receptor (EGFR)—were similarly enriched (*21*). The detection of supermeres in human plasma thus positions them alongside microvesicles, exosomes, and exomeres as promising targets for liquid biopsy diagnostics (*28*). This diagnostic potential is particularly relevant in the era of precision medicine, where the ability to distinguish subtle differences in biomarker profiles can directly impact patient-specific therapeutic strategies. However, harnessing their full diagnostic potential necessitates robust isolation and characterization methods, especially given their diminutive size and the overlapping biophysical properties with other nanoparticle fractions in biofluids (*22,25*).

The technical challenges associated with the characterization of supermeres and related extracellular nanoparticle subpopulations are considerable. Although the conventional differential ultracentrifugation technique (*21,22*) is effective for enriching these nanoparticles, it requires a large sample volume and days of serial ultracentrifugation steps. Additionally, the particles often co-sediment as heterogeneous mixtures containing other vesicles, protein aggregates, and other nanoparticles (*9,18,29*). This labor-intensive process not only increases the risk of cross-contamination but also compromises the sensitivity and specificity of downstream analyses as these nanoparticles carry overlapping biomarkers (*9,21*). Even with optimized high-speed spins (e.g., 167,000×g for exomeres followed by 367,000×g for supermeres) (*22*), the isolation of supermeres from the contamination by serum proteins and particles of similar dimensions remains challenging, Density gradient ultracentrifugation, using media such as iodixanol or sucrose, has improved purity by more effectively separating vesicles from smaller macromolecular complexes (*9*), while techniques like asymmetric flow field-flow fractionation have enabled the initial segregation of exomeres from classical EVs based on differences in hydrodynamic size and density (*30,31*). These technical enhancements are critical not only for a clearer understanding of supermere biology but also for translating these findings into actionable clinical insights. Emerging methodologies, including microfluidic sorting (*32-36*) and immunoaffinity capture (*37,38*), have been applied to isolate specific EV subsets; however, these approaches are constrained by limitations such as the need for prior knowledge of inertial properties, precise non-overlapping isoelectric point, specific surface markers, and scalability challenges in clinical settings. Consequently, there remains an unmet need for a selective, user-friendly platform capable of characterizing shared proteins on novel nanoparticles such as supermeres directly within complex biological samples.

To address this gap, we introduce the Ion Exchange Membrane Sensor (IEMS), a novel, charge-gated immunosensor based on permselective membranes that distinguishes supermeres label-free from other extracellular particles by integrating charge-based gating with marker-targeted capture. We used IEMS with silica nanoparticle labels in our previous studies to characterize exosomes, lipoproteins, and proteins (*15,16,24,39*), unlike the current label-free approach. The IEMS leverages a hydrophilic anion exchange membrane functionalized with antibodies against a selected protein marker, thereby enabling the comprehensive capture of all entities bearing this marker, including supermeres, other nanocarriers, and soluble proteins. It is a charge-based sensor that uses surface charge on the membrane to gate the ion current through the membrane. It employs the electroconvective instability of permselective membranes (in the overlimiting current region) to amplify the current signal of the surface charge. Critically, among nanoparticles, only supermeres as a nanocarrier exhibit a zeta potential of -50 mV (based on our measurements in this work from across multiple sources) that is significantly more negative than the thermal fluctuation threshold (∼ – *R*_*B*_*T/F* =-25 mV) while remaining substantially larger than soluble proteins and nucleic acids. Hence, the IEMS not only amplifies the signal from the charged supermeres but suppresses noise from the weakly charged interfering proteins in the sample. This unique combination of biophysical characteristics yields a measurable signal exclusively attributable to supermeres even in highly heterogeneous physiological samples like plasma—a selectivity that has been rigorously validated through appropriate control experiments. This sensitivity (10^6^-10^7^ copies/ml limit of detection) and selectivity features of the IEMS enable isolation-free and rapid (<30 minutes) supermere detection and quantification in untreated plasma over a 3-log dynamic range, thus overcoming the significant limitations of conventional days-long methods and permitting validation of supermere as a viable biomarker with extensive testing of clinical samples.

Complementary analyses using orthogonal techniques have confirmed that supermeres possess a notably negative charge (approximately –50 mV), attributable to the presence of RNA and an array of surface-accessible proteins—including ENO1, ENO2, HSPA13, DDR1, CEACAM5 (CEA), and TGFB induced (TGFBI)—while classical tetraspanins are undetectable. Importantly, markers such as HSPA13, ENO2, and DDR1 have demonstrated enhanced specificity for supermeres, effectively serving as analogues to tetraspanins in their diagnostic utility. Comparative diagnostic evaluations further revealed that supermeres outperform exomeres and small EVs in detecting colorectal cancer, as evident by a significant differential expression in a cohort of 22 colorectal cancer patients versus 25 healthy controls. Similar trends were observed in additional, albeit smaller, cancer cohorts, with each group exhibiting distinct overexpression profiles. Notably, in two colorectal cancer patients who underwent tumor resection, a marked postoperative decline was observed exclusively in supermere levels—a finding corroborated by ultracentrifugation coupled with surface plasmon resonance analyses. These results further underline the potential of supermere-focused diagnostics to overcome the pitfalls of traditional isolation methods, ultimately supporting more precise clinical decision-making.

Collectively, these findings underscore the potential of supermeres as a novel diagnostic target and highlight the utility of the IEMS platform for rapid, direct characterization of extracellular nanoparticles in clinical biofluids. By integrating a detailed understanding of the biophysical properties of supermeres along with a novel characterization platform, this work highlights supermeres as a promising biomarker class that not only fills a critical gap in our current diagnostic tools but also overcomes the limitations of conventional methodologies.

## 2. Results

### 2.1. Size and zeta potential of supermeres, exomeres, and sEVs

We isolated supermeres, exomeres, and small extracellular vesicles (sEVs) pellets from multiple cancer cell-culture media—including a fibroblast control—by differential ultracentrifugation (Fig. 1a) with multiple rounds of pelleting and resuspension to ensure higher purity (*22*). Particle size distributions were determined by dynamic light scattering (DLS), and the surface charge was quantified via electrophoretic light scattering (zeta potential). DLS revealed distinct modal diameters: supermeres (∼25 nm), exomeres (∼40 nm), and sEVs (∼100 nm) (Fig. 1b). Although DLS is prone to size overestimation, it reliably distinguishes relative differences in size. The particle peaks differ; however, the overlap among distributions and co-isolated soluble lipoproteins precludes robust size-based characterization alone, especially from plasma with high lipoprotein concentrations.

**Figure 1.**
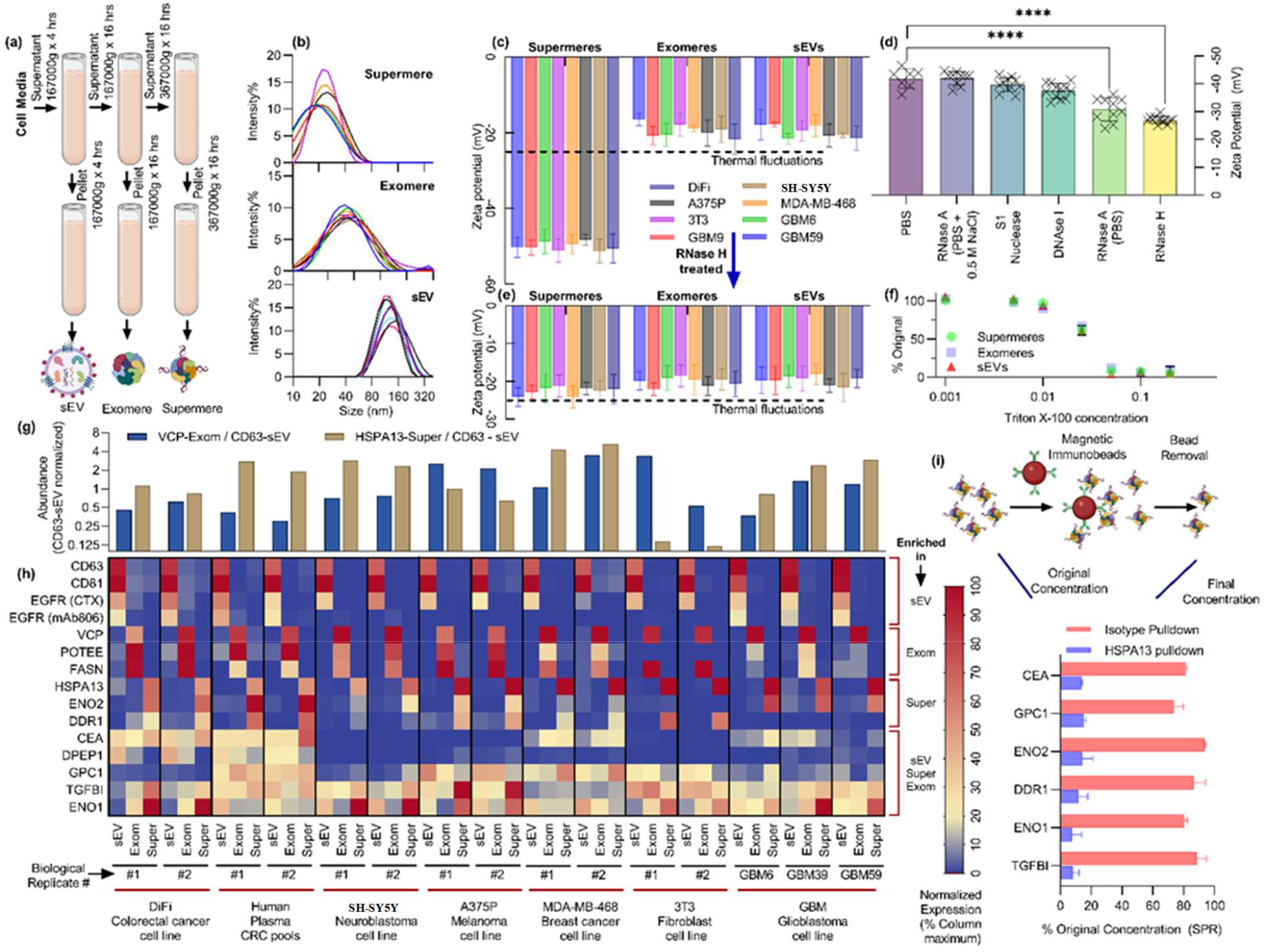
Characterization of supermeres, exomeres, and small extracellular vesicles (sEVs). (a) Schematic of sequential ultracentrifugation-based isolation yielding highly pure fractions of supermeres, exomeres, and sEVs from cell culture media and diverse biofluids. (b) Dynamic light scattering size distributions demonstrating discrete populations for supermeres (∼20–30 nm), exomeres (∼30–50 nm), and sEVs (∼100– 160 nm). (c) Zeta potential measurements across multiple cell lines reveal a significantly greater negative surface charge on supermeres compared with exomeres and sEVs. (d) Treatment of supermeres with different nucleases significantly reduced their negative zeta potential, confirming surface-associated RNA as the source of high charge. (e) Treatment with RNase H-induced zeta potential reduction in supermeres isolated from different cell lines. (f) Triton X-100 solubility assays indicate similar detergent sensitivity of supermeres, exomeres, and sEVs. (g) Surface plasmon resonance (SPR) quantification of HSPA13-positive supermeres and VCP-positive exomeres relative to CD63-positive sEVs. (h) SPR protein profiling highlights unique marker enrichment: HSPA13, DDR1, and ENO2 in supermeres; VCP, FASN, and POTEE in exomeres; CD63, CD81, and EGFR in sEVs; with CEA, DPEP1, GPC1, TGFBI, and ENO1 common to all fractions. (i) Immunoprecipitation of HSPA13 from supermere preparations markedly depleted DDR1, ENO2, CEA, GPC1, ENO1, and TGFBI from the supermere fraction but not with immunoprecipitation with isotype control, demonstrating their co-association with HSPA13 and supporting their designation as supermere-specific markers.

Furthermore, zeta potential measurements (Fig. 1c) showed that supermeres carry a significantly higher negative surface charge than exomeres or sEVs. The magnitude of the supermere zeta potential (>> 25 mV) exceeds the thermal noise threshold (*R*_*B*_*T/F* ≈25 mV), whereas exomeres and sEVs fall below it. As surface charge density scales exponentially with zeta potential, supermeres possess a net surface charge several orders of magnitude greater than that of exomeres or sEVs. The molecular basis for this elevated charge is examined in subsequent sections.

### 2.2 Supermeres carry RNAs on their surface and are solubilized in detergents

To elucidate the origin of the surface charges, supermeres were treated with various nucleases, followed by ultracentrifugation, buffer exchange, and resuspension under uniform initial conditions. These nucleases specifically target different types of nucleic acids, including single-stranded RNA (ssRNA), double-stranded RNA (dsRNA), single-stranded DNA (ssDNA), and double-stranded DNA (dsDNA). As shown in Fig. 1d, nuclease-mediated hydrolysis of surface-bound nucleic acids resulted in a reduction in the magnitude of the zeta potential. Our findings indicate that the negative surface charge on supermeres primarily arises from the presence of RNAs. The change in zeta potential was observed only in supermeres across all tested cell lines, with no effects seen on exomeres or sEVs after RNase H treatment (Fig. 1e). This observation aligns well with a previous report by Zhang *et al*., which also suggests RNAs are enriched in supermeres (*21*). A more detailed investigation is required to identify the specific type(s) of nucleic acid involved, which we plan to pursue in future studies.

We next assessed detergent solubility using a detergent solubility assay across varying concentrations. Briefly, nanocarrier fractions were treated with varying detergent concentrations, resuspended in PBS, and subsequently analyzed by surface plasmon resonance (SPR), as illustrated in Fig. 1f. Upon detergent-mediated solubilization of nanocarriers, target proteins bind individually to the SPR surface rather than as part of intact nanocarriers, resulting in decreased SPR signals compared to intact nanocarrier binding. Supermeres showed solubility in Triton X-100 at approximately 0.05–0.1%, based on the SPR signal pre- and post-treatment. This behavior was similar to that of exomeres and sEVs, despite supermeres lacking a clearly defined lipid mono- or bilayer enclosure. The observed solubility of supermeres suggests that detergent molecules, with their hydrophobic and hydrophilic domains, can effectively disrupt the interactions that stabilize supermeres, similar to the behavior observed in exomeres and sEVs. This solubility property further distinguishes supermeres from amyloid-like aggregates, which typically resist detergent solubilization or require substantially higher detergent concentrations.

### 2.3. Protein distribution of supermeres and other nanocarriers

We next examined the protein composition in supermeres, exomeres, and sEVs derived from various cell cultures using SPR, using higher purity nanocarrier fractions (multiple resuspensions/pelleting), specifically targeting major proteins on different nanocarriers, including TGFBI, ENO1, GPC1, CEA (CEACAM5), DPEP1, HSPA13, DDR1, ENO2, FASN, VCP, POTEE, CD9, CD81, CD63, and EGFR. These proteins were selected based on prior studies identifying their prominence and functional relevance within these particle types (*21,22,26*). The SPR data (Fig. 1g,h) indicate that the tetraspanins CD63 and CD81, and EGFR were mostly absent from supermeres and exomeres but highly enriched in sEVs. Conversely, FASN, VCP, and POTEE were primarily associated with exomeres. Proteins HSPA13, DDR1, and ENO2 were specific to supermeres, showing minimal presence in other fractions across different cell culture media. Additionally, TGFBI, ENO1, GPC1, CEA, and DPEP1 appeared significantly across all three fractions. Collectively, the protein distribution results (Fig. 1g,h), supported by DLS and zeta potential findings, substantiate that supermeres represent a distinct nanocarrier population compared to exomeres and sEVs. Also noteworthy is that while HSPA13+ supermeres were expressed significantly relative to CD63+ sEVs for all cell lines studied, they were significantly less abundant in the healthy fibroblast control (Fig. 1g), and this is something that is also shown for healthy human subjects in subsequent sections. Moreover, different proteins seem to be more abundant on supermeres for some cell lines than others. This could potentially pave the way for disease-type detection in our future work, with some limited data shown in a subsequent section with actual human samples.

The SPR data also suggests that HSPA13, DDR1, and ENO2 markers are present on a large percentage of supermeres in a similar way that tetraspanins are found on many EV populations (Fig. 1h). To validate this, we conducted HSPA13 immunoprecipitation on isolated supermere fractions, assessing the coprecipitation of other supermere-associated proteins relative to isotype control (Fig. 1i). The results confirm HSPA13’s extensive colocalization with multiple supermere proteins, suggesting that supermeres could potentially be isolated using immunoseparation methods targeting HSPA13, DDR1, or ENO2. However, the significantly lower abundance of these proteins in isolated supermere fractions relative to non-specific proteins, along with the low immunoseparation yields, suggests a high degree of supermere heterogeneity, posing substantial challenges for their characterization. Thus, while immunoseparation is feasible for isolating specific supermere subpopulations based on unique surface proteins, it will be less selective for proteins shared with other nanocarriers, such as CEA, TGFBI, and ENO1. Our objective, therefore, is to develop robust and direct characterization methods capable of efficiently analyzing any supermere subfraction in untreated samples within a practical timeframe, including proteins shared with exomeres and sEVs.

### 2.4. Direct quantification from complex biofluids in <30 minutes using distinctly different zeta potential of supermeres

To further characterize supermeres, we utilized a novel Ion Exchange Membrane Sensor (IEMS) platform that distinguishes analytes captured on its surface based on their inherent charge—or the charge on their reporter nanoparticles when a sandwich scheme is employed. This method was previously validated for lipoprotein, protein, and sEV characterization (*15,16,24,39*); however, for these analytes, a charged silica reporter was necessary due to their negligible native surface charge. It is important to note that, in the cases of lipoproteins, proteins, and sEVs, no significant signal was observed without the charged silica reporter. We exploit this phenomenon to characterize supermeres: by targeting a protein that is shared across different nanocarriers, only supermeres will generate a signal owing to their substantially higher surface charge, while other nanocarriers will not produce a detectable signal.

The IEMS incorporates an anion exchange membrane (AEM), which allows the selective passage of anions under an applied electric field. Three distinct regimes characterize the current-voltage behavior of this system: the ohmic regime at lower voltages, the limiting regime at intermediate voltages, and the overlimiting regime at higher voltages (*15,16,24,34,39-43*). At lower voltages, the system follows Ohm’s law of the bulk electrolyte, as the voltage drop (resistance) is mostly in the bulk solution, such that the current increases linearly with respect to the voltage. With increasing voltage, the depletion of ions by the permselective membrane creates an ion-depleted region on one side of the membrane whose dimension is roughly that of the sensor (∼100 microns). Most of the voltage drop now occurs within this small ion-depleted region, and the ion current saturates at a constant asymptote with respect to increasing voltage in this “limiting-current” regime. At even higher voltages, an electroconvective instability introduces convective vortex mixing to replenish ions into the ion-depleted region, thus eliminating the asymptotic limiting current to produce a second rise in the current with respect to the voltage in the overlimiting regime. Without Debye screening under the ion-depleted conditions introduced by the permselective membranes, these transitions, particularly the onset of the electroconvective instability to initiate the overlimiting current, are highly sensitive to the membrane’s surface charge, which gates the ion current into the permselective membrane and controls the tangential surface electric field that drives the electroconvective instability. The IEMS exploits these surface charge sensitivities of the IEMS to directly characterize highly charged supermeres in biofluids.

Fig. 2a presents a schematic of the IEMS platform, while Fig. 2b illustrates a representative current–voltage response. To ensure accurate quantification across experiments, the measured signal was normalized to account for membrane size variations and limiting current (Figure S1, Supporting Information), yielding a charge-based signal in femto Coulombs (fC). Validation with isolated and SPR quantified supermeres from DiFi cell media established a detection limit of approximately 3×10^6^ supermeres/mL (∼5 fM) and sensor saturation at about 10^9^ supermeres/mL (∼10 pM), achieving a dynamic detection range spanning over three orders of magnitude for all surface markers considered (Fig. 2c).

**Figure 2.**
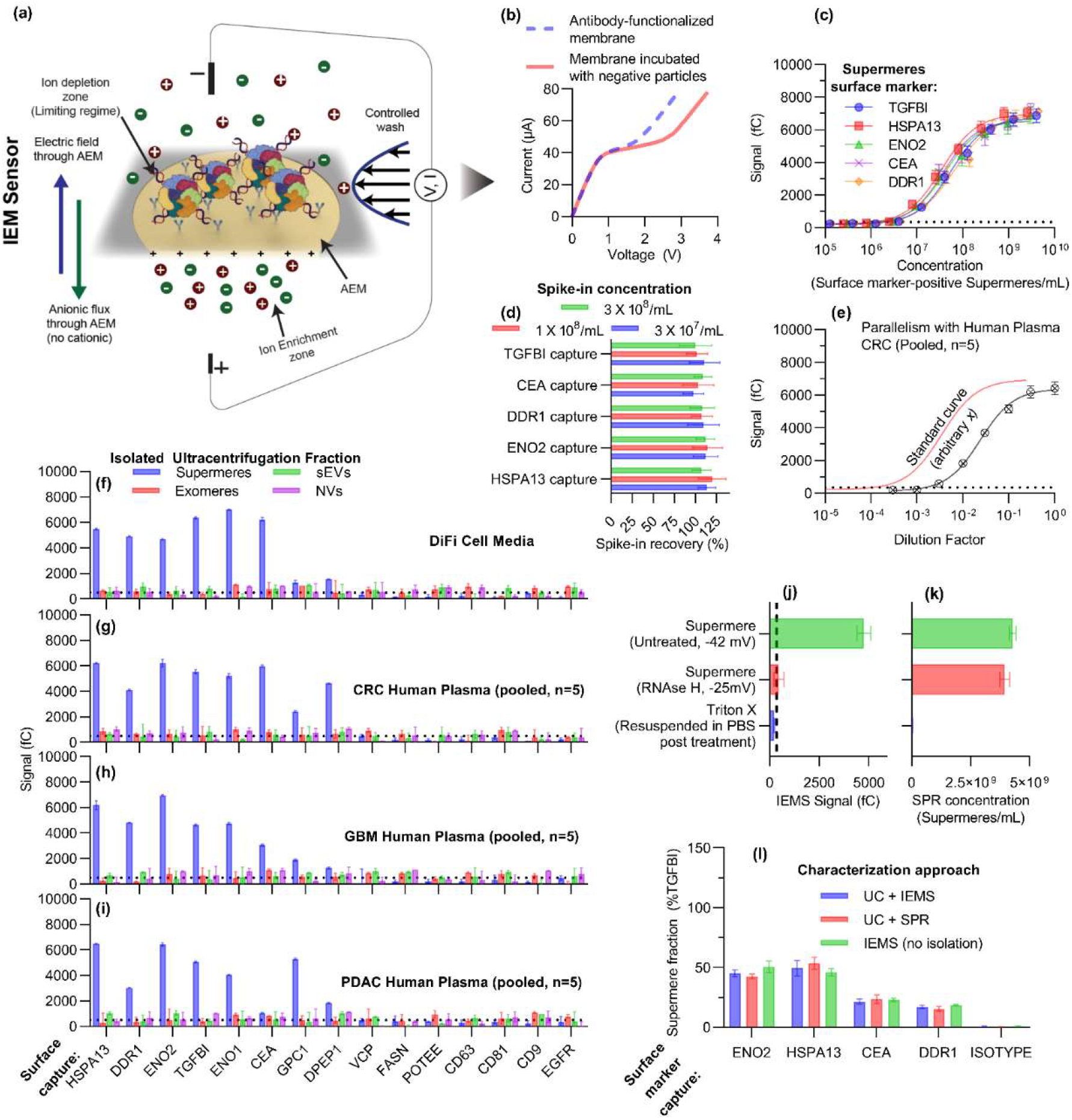
Rapid and direct characterization of supermeres using the Ion Exchange Membrane Sensor (IEMS) (a) Typical schematic of IEMS for supermere characterization with antibodies functionalized on its surface (b) Typical current-voltage curve for IEMS. (c) Calibration plot surface marker-positive supermeres measuring IEMS signal against the known concentration of supermeres (from SPR). (d) Spike-in recovery experiments in healthy plasma show near-complete recovery of DiFi-derived supermeres across low to high concentrations, underscoring the sensor’s accuracy and reproducibility in complex biofluids. (e) Parallelism assessment for HSPA13-positive supermeres, wherein a pooled colorectal cancer (CRC) plasma sample is serially diluted and yields a curve parallel to the standard curve generated from isolated DiFi supermeres, indicating minimal matrix interference. (f–i) Comparison of IEMS signals for supermeres, exomeres, small EVs (sEVs), and the non-vesicular fraction (post-filtration) isolated from (f) DiFi cell media or (g) pooled plasma from CRC, (h) glioblastoma GBM, and (i) pancreatic adenocarcinoma PDAC patients. Only supermeres generate a charge-based signal—regardless of marker—while exomeres, sEVs, and the non-vesicular fraction remain undetected. (j,k) Effect of RNase H and detergent on signal of SPR and IEMS (k) showing charge colocalization with surface markers on supermeres. (l) Direct quantification of supermeres from cell media using IEMS, compared to ultracentrifugation (UC) followed by SPR or IEMS, shows excellent agreement across all three methods, validating the rapid (<30 min) and isolation-free capabilities of the IEMS platform.

Critically, the platform demonstrated great analytical performance in both spike-in recovery and parallelism assessments. Spike-in recovery experiments, were conducted by adding UC-isolated supermeres from DiFi media into a pooled healthy human plasma sample (our study suggests the healthy human plasma does not carry any significant amount of TGFBI+, CEA+ DDR1+, ENO2+, and HSPA13+ supermeres as discussed in next section, see Figure 3), resulted in nearly 100% recovery across low, mid, and high concentrations within the dynamic range (Fig. 2d). This high recovery rate underscores the assay’s accuracy and reproducibility in complex biological matrices. Moreover, when a pooled colorectal cancer (CRC) patient plasma sample was serially diluted with measured signal as shown in (Fig. 2e) for HSPA13+ supermeres, it was parallel to the standard curve obtained for isolated DiFi supermeres, showing parallelism. The idea behind parallelism is to show that if any inhibitor or interfering species were present in the plasma that interferes with our sensor signal, it gets serially diluted and thus should deviate from being parallel to a standard curve generated using isolated supermeres. The spike-in recovery and parallelism demonstrate the robust and sensitive nature of our platform.

**Figure 3.**
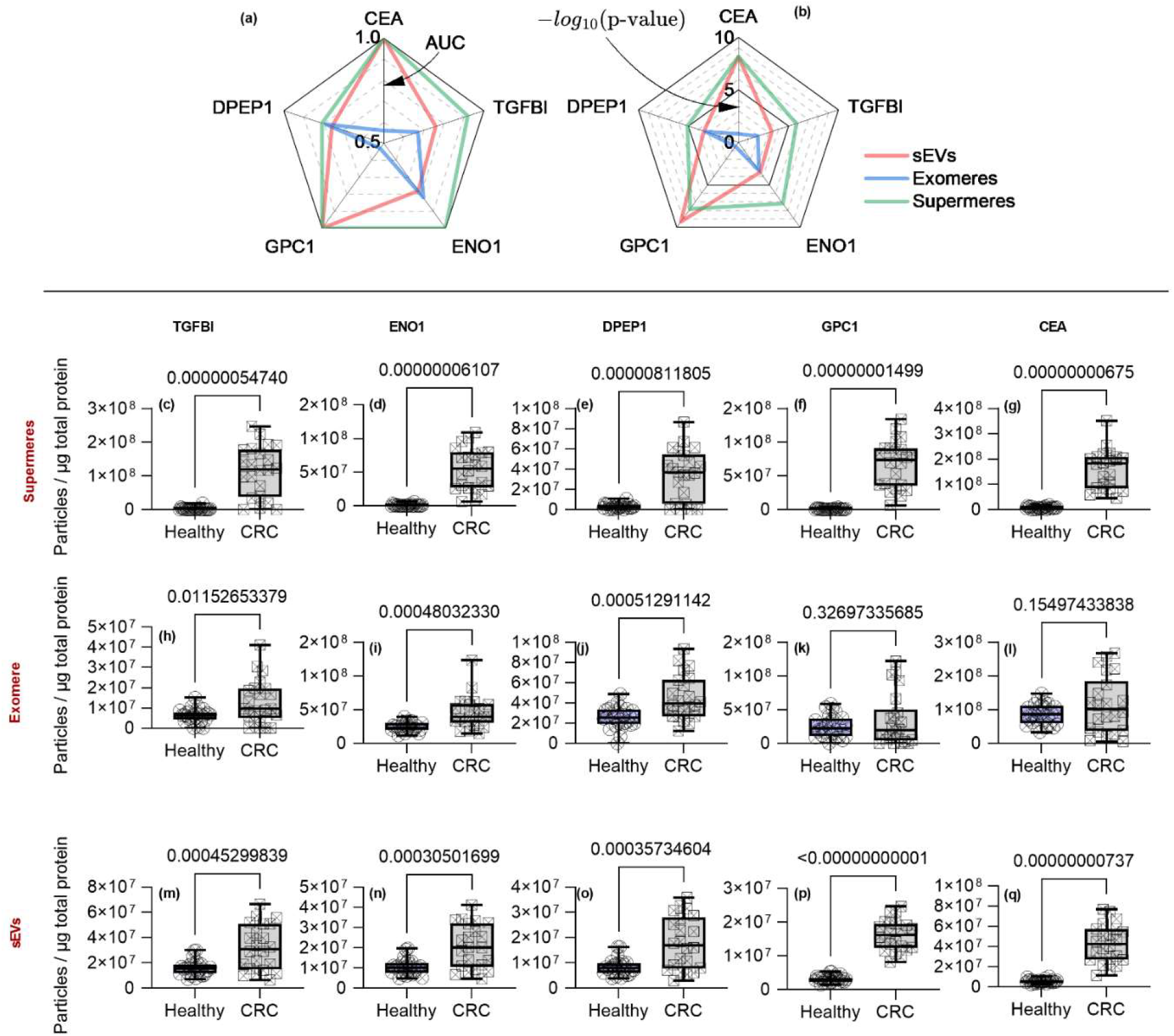
Diagnostic comparison of supermeres, exomeres, and sEVs in colorectal cancer (CRC). The use of different proteins on different nanocarriers in diagnosing a cohort of healthy and colorectal cancer patients. (a) and (b) show the AUC and p-values, respectively, for each of the proteins on different nanocarriers, showing supermeres outperform the other nanocarriers. Supermeres (c-g), Exomere (h-l), and sEVs (m-q) were characterized with TGFBI (c,h,m), ENO1 (d,i,n), DPEP1 (e,j,o), GPC1 (f,k,p), and CEA (g,l,q).

Next, we analyzed DiFi cell media and pooled human plasma samples from colorectal cancer (n=5), glioblastoma (n=5), and pancreatic adenocarcinoma (n=5) samples. We isolated —supermeres, exomeres, sEVs—and a final non-vesicular fraction (filtered to remove entities larger than 300 kDa), nanocarrier fractions using ultracentrifuged material, and subsequently evaluated the signal produced by each antibody-captured supermere under investigation using IEMS. As shown in Fig. 2f–I, the signal was exclusively produced in the supermere fraction and only when the probe specifically targeted a protein that is present on supermeres. For proteins that are shared with other nanocarrier fractions, no signal was detected in any fraction other than supermeres, regardless of whether the source was DiFi media or plasma from different disease groups. Even TGFBI, CEA, and tetraspanins, which are abundant in exomeres/sEVs, do not produce a signal in those fractions, as these nanocarriers are weakly charged and IEMS only produces signals with charged molecules or particles.

Furthermore, to demonstrate that the charge detected by our sensor is indeed co-localized with other proteins on supermeres, we compared untreated supermeres with those treated with RNase H and detergent using both SPR and IEMS. As illustrated in Fig. 2j and 2k, we observed that SPR produced a signal with both untreated and RNase H-treated supermeres, indicating that the nanocarrier structure remains intact following nuclease treatment. In contrast, IEMS did not produce any signal with RNase H-treated supermeres, as the charge-carrying species on the supermeres had been hydrolyzed. Notably, both SPR and IEMS utilize the same capture probes; hence, both techniques successfully captured the supermeres, but the absence of charge in the RNase H-treated sample prevented IEMS from generating a signal. This outcome confirms the co-localization of the charge with the proteins on supermeres. Additionally, detergent-treated supermeres failed to produce any signal in both SPR and IEMS analyses due to the loss of structural integrity of supermere proteins, as the detergent solubilized the supermeres, thereby eliminating both the protein epitopes and the charge required for detection. It is important to note that among all the treatments, a final buffer exchange was performed to remove any buffer-based effect on SPR results or antibody-antigen kinetics.

Direct characterization from biofluids was demonstrated by characterizing UC-isolated DiFi supermeres with both SPR and IEMS and compared to direct characterization of supermeres without ultracentrifugation using IEMS as shown in Fig. 2l – all three methods showed identical results. The concentrations were normalized with respect to TGFBI supermeres to account for any yield bias during ultracentrifugation. Results from all three approaches showed identical outcomes (Fig. 2l), underscoring the sensitivity, robustness, and specificity of the IEMS method, corroborated by the orthogonal SPR technique. More importantly, it underscores that the more than 2 days long characterization with UC + SPR or UC + IEMS produced identical results compared to less than 30 minutes of characterization using IEMS without any isolation.

### 2.5. Supermeres demonstrate superior or comparable CRC diagnostic performance to sEVs, while exomeres exhibit limited efficacy via UC + SPR

Using UC+SPR as a consistent method for characterizing all three nanocarrier fractions—supermeres, exomeres, and sEVs—we evaluated their effectiveness in distinguishing colorectal cancer (CRC) patients from healthy individuals (Fig. 3). We focused on the proteins TGFBI, CEA, ENO1, DPEP1, and GPC1, which are present across all three particle types (Fig. 1h). The spider plots (Fig. 3a,b) demonstrate that supermeres exhibited better or comparable Area Under the Curve (AUC) values and p-values compared to sEVs and notably outperformed exomeres which had weak or no diagnostic potential. While GPC1 showed equivalent diagnostic performance in supermeres and sEVs, supermeres generally provided enhanced diagnostic performance for the remaining proteins. Exomeres, however, demonstrated limited diagnostic capability, with significantly lower discriminative power between CRC patients and healthy individuals. Protein expression levels in healthy controls were markedly higher in sEVs and exomeres, while supermeres were predominantly absent or minimally present (Fig. 3c-q). Furthermore, proteins like GPC1, although abundant in exomeres, offered minimal diagnostic relevance in this fraction.

Subsequently, we analyzed proteins uniquely associated with each UC-isolated nanocarrier type: HSPA13, ENO2, and DDR1 for supermeres; VCP, POTEE, and FASN for exomeres; and CD63, CD81, and EGFR for sEVs. These proteins collectively reflect the overall presence of their respective nanocarriers in plasma. Fig. 4a-i illustrates that supermere-specific proteins were largely absent in healthy controls, whereas exomere- and sEV-specific proteins did not demonstrate significant differences between CRC patients and healthy individuals. Notably, supermeres showed significantly better diagnostic performance based on AUC and p-values for their associated proteins, suggesting a potential direct involvement in cancer pathogenesis. All evaluated supermere-associated proteins showed excellent performance in diagnosing CRC patients, underscoring their clinical potential.

**Figure 4.**
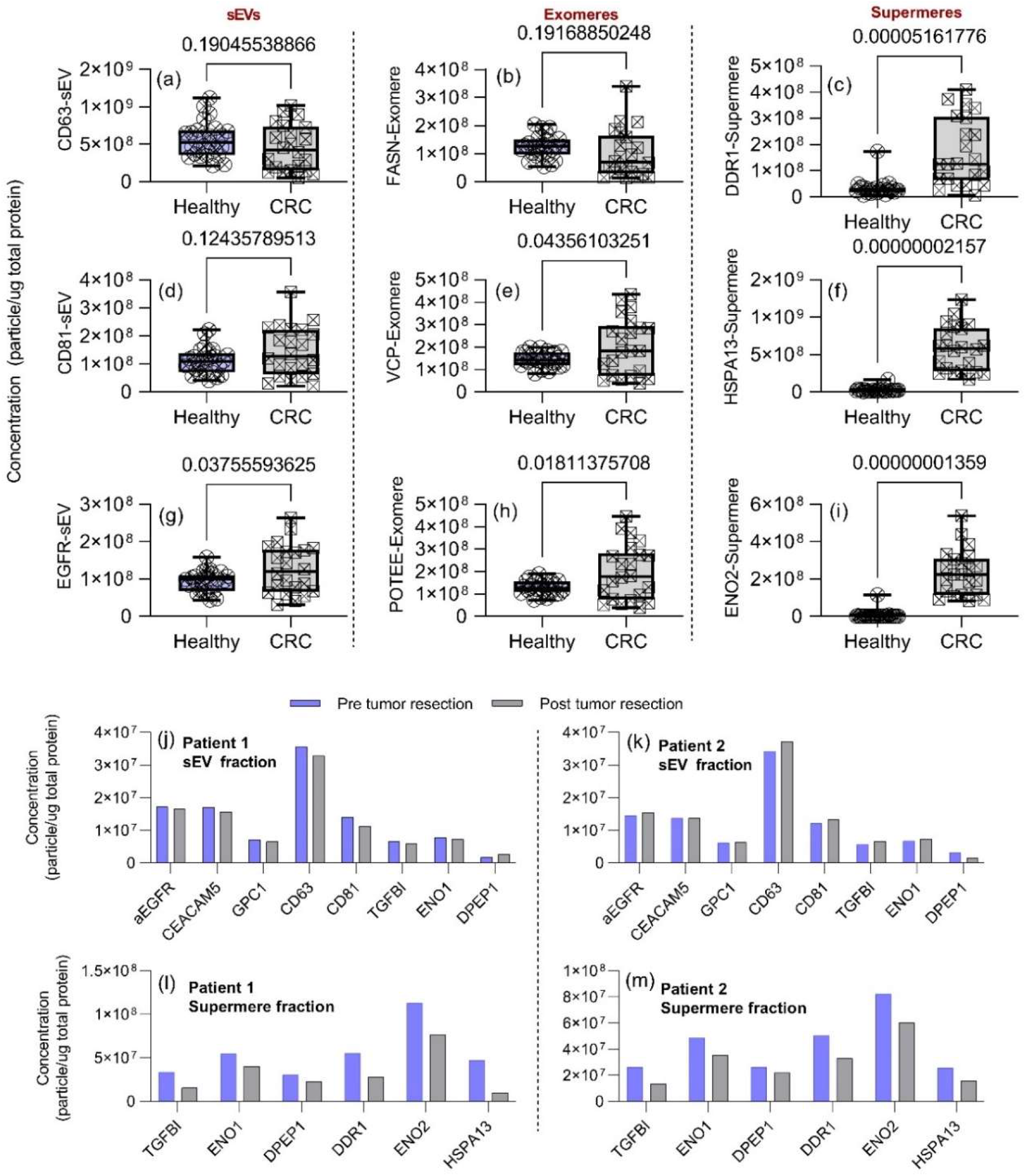
Comparative analysis of sEV-, exomeres-, and supermere-specific proteins in healthy versus colorectal cancer (CRC) plasma, and their changes following tumor resection. (a–i) Box-and-whisker plots illustrating the plasma concentrations of selected proteins associated with each nanocarrier subtype—small extracellular vesicles (sEVs), exomeres, and supermeres—in healthy controls (left) versus CRC patients (right). Notably, supermere-specific markers as shown in (c), (f), and (i) (DDR1, HSPA13, ENO2) are significantly elevated in CRC patients compared to healthy individuals, whereas exomere- and sEV-specific markers show less pronounced discrimination. (j, k) Concentrations of key proteins on sEVs and (l, m) supermeres in two CRC patients pre- and post-tumor resection. Substantial decreases in supermere-associated proteins are observed post-surgery, whereas sEV-associated proteins exhibit relatively minor changes. These findings underscore the superior diagnostic utility of supermeres over conventional EV markers.

Additionally, we analyzed plasma samples from CRC patients collected before and after tumor resection surgery. Post-surgical samples showed a substantial reduction in supermere-associated proteins, while minimal or negligible changes were observed in the sEV-associated proteins (Fig. 4j-m). These observations highlight the clinical relevance of supermeres and suggest their potential utility in companion diagnostics, warranting further detailed investigation similar to previously studied sEVs.

The biological implications of these findings are significant. Supermeres were only recently identified as a distinct class of extracellular nanoparticles, and emerging evidence suggests they play active roles in disease processes (*26*). For example, cancer-derived supermeres can transfer aggressive traits such as cetuximab drug resistance and enhanced aerobic glycolysis (Warburg effect) to recipient cells, setting them apart as functional messengers rather than inert carriers. Moreover, supermeres exhibit markedly greater biodistribution and tissue uptake in vivo compared to extracellular vesicles, and they harbor a large proportion of extracellular RNA released by tumors, suggesting that tumors shed a prolific number of supermeres loaded with disease-relevant cargo. This could explain why our readouts for the supermere fraction are so distinct—we are essentially capturing a concentrated stream of tumor-derived information. By contrast, the sEV and exomere fractions contain more “mixed” content (generic vesicle markers or metabolic proteins not exclusively tumor-specific), which may dilute their diagnostic signal relative to supermeres. These results highlight the value of parsing extracellular particles into subpopulations: by attributing specific biomarkers to their correct carrier (exosome, exomere, or supermere), we can identify which compartment is the richest source of cancer-derived signals. Here, the supermere fraction stands out as a rich and reliable source of cancer biomarkers, many of which were previously unrecognized or conflated within bulk EV analyses.

### 2.6. IEMS rapidly identifies supermeres in healthy vs CRC diagnosis with similar performance to UC + SPR

Although UC + SPR in the previous section utilized similar characterization methods across different isolated nanocarriers (Fig. 3 and 4), this section focused on IEMS performance in the characterization of supermeres in healthy individuals’ vs CRC patient plasma in less than 30 minutes. Samples were compared to PDAC (Pancreatic adenocarcinoma, n=5) and GBM (Glioblastoma, n=5) individual human plasma samples. GBM and PDAC samples picked here are different than those used previously in the pooled samples in Fig. 2. We chose HSPA13-positive supermeres due to their abundant nature across cancer types (Fig. 1h), CEA-positive supermeres because CEA is a gastrointestinal-specific marker, and ENO2-positive supermeres because of their abundance in neuronal and gastrointestinal tissues.

Using IEMS, we observe identical performance of HSPA13+, CEA+, and ENO2+ supermeres in healthy vs CRC, as shown in Fig. 5a-e, respectively, with them performing similarly to their counterparts in Fig. 4 using UC + SPR with similar AUC values. The study shows the variation of surface protein across different diseases, with HSPA13 supermeres abundant across the disease types parallel to their prevalence across different cell media, but ENO2 supermeres are only abundant in GBM and CRC due to their specific neuronal and gastrointestinal cancer expression. CEA supermeres were mostly only overexpressed in CRC compared to GBM, PDAC, and healthy human subjects making it most optimal at identifying CRCs. Total CEA in human plasma (measured using ELISA), on the other hand, was less efficient at differentiating healthy from CRC samples, with an AUC of 0.8575 (Fig. 5e).

**Figure 5.**
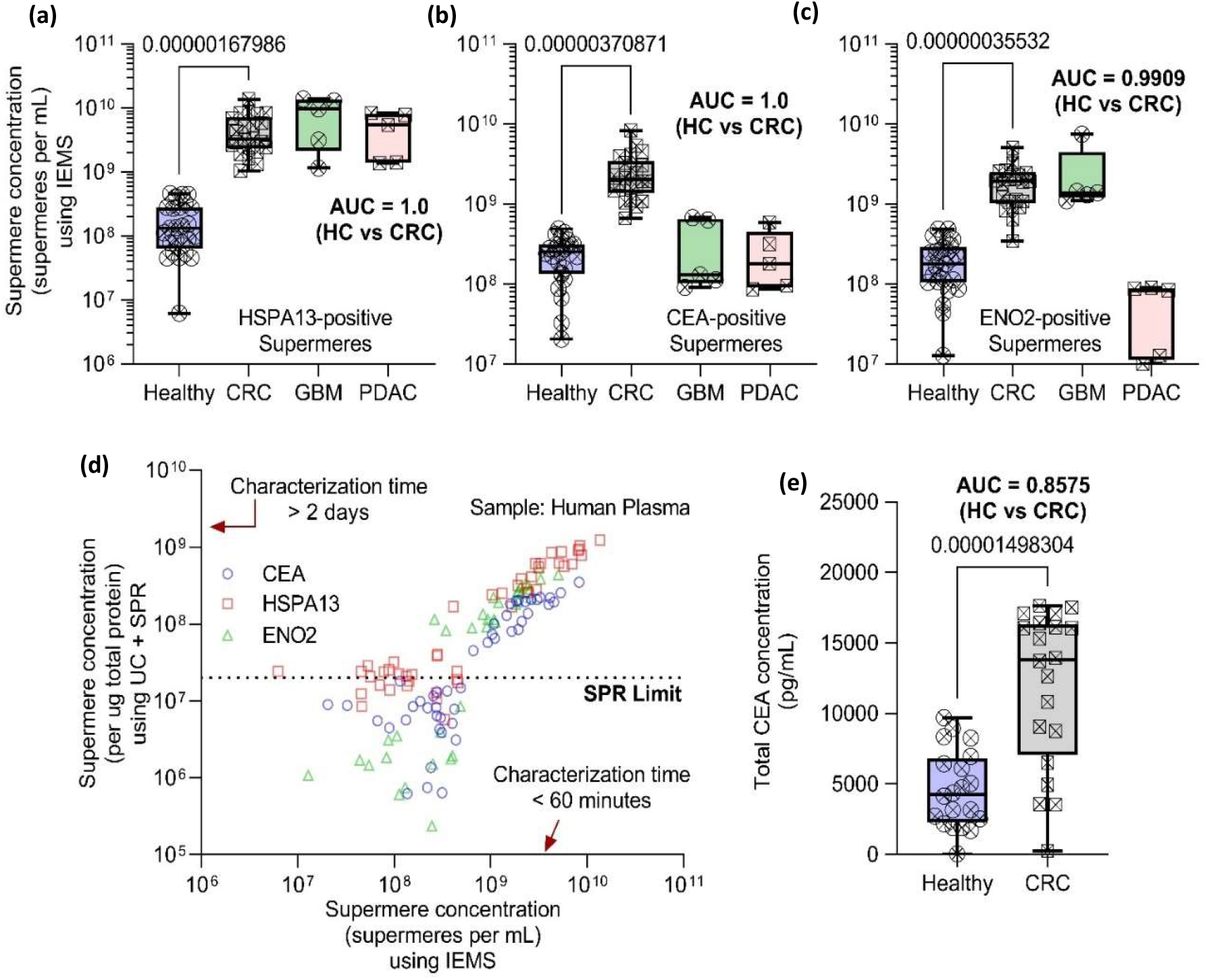
Rapid IEMS-based detection of supermeres in human plasma outperforms total CEA for colorectal cancer (CRC) diagnosis and correlates with orthogonal methods. (a–c) Box plots show significantly elevated concentrations of HSPA13-, CEA-, and ENO2-positive supermeres in CRC patients (pink) compared to healthy controls (blue), with additional data from glioblastoma (GBM, green) and pancreatic adenocarcinoma (PDAC, teal) cohorts. (d) Comparison of supermere concentrations measured by Ion Exchange Membrane Sensor (IEMS) versus ultracentrifugation plus Surface Plasmon Resonance (UC+SPR) shows a strong linear correlation across a range of concentrations. (e) Total CEA levels in plasma, measured by conventional ELISA, exhibit lower discriminatory power (AUC = 0.8575) relative to supermere-based markers (a–c). Box plots represent median, 25th–75th percentile (box), and 1.5× interquartile range (whiskers). Statistical analyses and sample sizes are indicated in the text. These results confirm that IEMS enables rapid (<30 min), sensitive quantification of supermeres, yielding superior or comparable CRC diagnostic performance to conventional biomarkers and multi-day UC+SPR methods.

Importantly, IEMS also detected small amounts of supermeres in healthy human plasma that were typically missed with SPR due to poor sensitivity of SPR (LOD ∼ 1e7 to 1e8 supermeres/mL) compared to IEMS (<1e6 to 1e7 supermeres/mL). Both IEMS and UC+SPR showed a linear trend in concentration above the limit of detection of SPR (Fig. 5d), demonstrating consistency between the orthogonal characterization method and across different surface capture types.

## 3. Discussion

Our study identifies and characterizes supermeres as a distinct class of extracellular nanoparticles that exhibit unique biophysical and molecular features compared to classical exosomes and exomeres. Supermeres display a significantly more negative zeta potential—and harbor a specialized protein repertoire, including markers such as HSPA13, ENO2, and DDR1. These features not only distinguish supermeres from other extracellular vesicle subtypes but also underscore their potential as precise and robust biomarkers for cancer diagnostics.

By leveraging the novel Ion Exchange Membrane Sensor (IEMS), we achieved rapid (<30 minutes) and sensitive direct quantification of supermeres from complex biofluids made possible by the charge characteristics of supermeres. The IEMS platform effectively overcomes the limitations of conventional isolation methods, demonstrating an extensive dynamic detection range, high recovery, and reproducibility—features validated through parallel assessments with ultracentrifugation coupled with Surface Plasmon Resonance (UC + SPR) that takes over two days for characterization. Notably, our comparative analyses reveal that supermeres provide superior diagnostic performance compared to small extracellular vesicles and exomeres in distinguishing CRC patients from healthy individuals, and the distinct expression profiles of supermere-associated proteins across different cancer types highlight their potential for disease-specific detection.

Overall, this work not only deepens our understanding of extracellular nanoparticle heterogeneity but also sets the stage for integrating supermere-based diagnostics into precision medicine. Future studies aimed at expanding validation in larger and more diverse patient cohorts will be critical for translating these findings into clinical practice. Studies understanding the biogenesis of supermeres can be very important in understanding their biological context and importance.

## 4. Experimental Section

### Ethics Statement and Human Plasma Samples

We received colorectal cancer and control group samples from Precision for Medicine and Vanderbilt University School of Medicine, Nashville. An approved IRB protocol is already in place at Precision for the collection and at Vanderbilt University School of Medicine for the collection of plasma samples from patients. Both work with regulatory authorities and accrediting organizations around the world to ensure that the sample collection process and protocol follow the latest FDA, EMA, and MHRA guidelines. The pooled healthy plasma samples were obtained from Innovative Research (IPLAK2E10ML).

### Fabrication of anion-exchange membrane (AEM) sensor

The anion exchange membrane (AEM) composed of polystyrene-divinylbenzene fine particles with strong basic quaternary ammonium groups (R-(CH_3_)_3_N^+^) supported by polyethylene as a binder and polyamide/polyester textile fiber, was obtained from Mega a.s. (Straz pod Ralskem, Czech Republic) and used as a sensor. The AEM sensor was fabricated by embedding a small piece of anion exchange membrane of approximate dimensions of 0.3 mm × 0.9 mm in an epoxy resin (TAP Quik-Cast, Tap plastic) using an optimized reported protocol^28^. Briefly, the membrane was hand-cut and placed on the tip of a silicone mold. A glass slide was then placed on top of the membrane piece. A two-component (side A and side B, 1:1 ratio) epoxy polyurethane resin mixture was then pushed to embed the AEM sensor. A 3D-printed sensor reservoir was then attached to the sensor disc using the same polyurethane resin mixture. The prepared sensor was soaked in de-ionized (DI) water overnight before further use.

### Fabrication of biochip

A 25 mm × 54 mm (w × l) size biochip having a microfluidic channel of 3 mm × 35 mm × 250µm (w × l × h) was fabricated. Three layers of a polycarbonate sheet of 0.3 mm thickness were used to fabricate the chip using our reported protocol^28^. Briefly, the sheets with orifices for the inlet, outlet, sensor, and channel were cut using a cutting plotter (FC700, Graphtec Corp., Japan). The sheets were then thermally bonded in an oven (Fisher Scientific, Isotemp Oven) at 177 °C for 15 min. The small tubes for the inlet and outlet, and three different-sized tubes were attached between the inlet and outlet for mounting a sensor and different electrodes using Acrifix UV glue. The electrode reservoirs were filled with 2% agarose gel to create a barrier between the microfluidic channel and the reservoirs.

### Functionalization of the antibody on the AEM surface

The surface modification of the anion-exchange membrane and antibody functionalization followed a previously optimized protocol^28^. Initially, the membrane surface was treated with a 0.1M solution of 3,3’,4,4’-Benzophenonetetracarboxylic acid (Sigma-Aldrich, USA) at pH 7 for 10 minutes. Subsequently, the surface was exposed to UV light at 365 nm (using an Intelli Ray 600 shuttered UV floodlight) for 90 seconds while purging with N2 gas. The sensor was then rinsed with deionized (DI) water. This surface modification process was repeated three times to ensure the generation of an adequate number of –COOH groups on the membrane surface. Following that, the sensors were immersed in DI water at pH 2 for 5 hours and subsequently washed with 0.1X PBS at pH 7. The carboxylated membrane surface was then ready for functionalization with the antibody probe using EDC (Thermo Fisher, USA) coupling chemistry. A 20 µL solution of 0.4 M EDC in 50 mM MES buffer at pH 6 was applied to the sensor surface for 40 minutes. The solution was then removed, and the sensor was washed with 1X PBS. Finally, a solution of the probe antibody (0.1 mg/mL) was incubated overnight on the sensor surface at 4ºC.

### Ultracentrifugation

The isolation and quantification of small extracellular vesicles (sEVs) from human plasma and cell media involved a detailed ultracentrifugation protocol. Initially, 200 µL of human plasma is diluted with 1xPBS to a final volume of 1 mL, or 1 mL of concentrated cell media is used directly (concentrated using a 100kDa filter from 10 mL to 1 mL). This mixture is centrifuged at 12,000 g for 20 minutes to remove larger particles. The supernatant is then passed through a 220 nm filter to eliminate larger debris. The filtered fluid is added on top of 3 mL of 1xPBS in a 4 mL ultracentrifuge tube and ultracentrifuged at 167,000 x g for 1.5 hours using a swinging bucket rotor (Beckman Coulter SW60Ti), which is preferred for its efficiency in pelleting vesicles compared to fixed angle rotors. After ultracentrifugation, the supernatant is carefully removed, leaving about 0.2 mL to avoid disturbing the pellet, which is then resuspended in 0.5 mL of ice-cold 1xPBS. For further purification, this resuspended solution is transferred on top of 15.5 mL of 1xPBS in a 17 mL ultracentrifuge tube and ultracentrifuged at 167,000 x g for 4.5 hours (SW32.1Ti). This step aims to refine the sEVs by pelleting them again under high-speed centrifugation. After pelleting, the sEVs are resuspended and passed through a 300 kDa filter to remove soluble proteins, thus enhancing the purity of the sEV sample. This additional purification step ensures the isolation of high-quality sEVs, now ready for downstream analyses such as surface plasmon resonance (SPR) to study their composition or concentration.

Exomeres and supermeres particles are isolated from the sEV-isolated supernatant using a previously reported protocol^22^. Briefly, the sEV-isolated supernatant is transferred to a new 4 mL ultracentrifuged tube and centrifuged at 167000xg for 6 hours in a swinging bucket rotor (Bechman Coulter SW60Ti) to pellet exomeres. The resulting exomere pellet is resuspended in ice-cold 1x PBS and further purified by ultracentrifuging at 167000xg for 18 hours. Meanwhile, the supernatant from the exomere spin is ultracentrifuged at 367000xg for 24 hours to isolate supermere particles. The resulting supermere pellet is further purified by an additional ultracentrifugation step at 367000xg for 24 hours. Finally, both exomere and supermere pellets are resuspended in ice-cold 1x PBS and passed through a 300 kDa filter to remove soluble proteins, thereby further enhancing the purity of the isolated samples.

### Surface Plasmon Resonance-based characterization of isolated sEVs

Before experimentation, all instrument-specific pre-experimental protocols recommended by the surface plasmon resonance (SPR) instrument manufacturer were followed, particularly those pertaining to “the maintenance chip” section. The system was then allowed to operate for an additional 12 hours using double-distilled water (DDI) in standby mode. Subsequent to this preparatory phase, the chip (Series S CM5 SPR chips, Cytiva, Catalog no. 29149603) was docked and normalized using 70% glycerol, or as otherwise advised by the manufacturer. The running buffer was then switched to phosphate-buffered saline (PBS) and maintained for two hours. Antibodies were buffer exchanged and resuspended in 10 mM MES buffer at a pH of 6.0. This solution was flowed over the SPR chip at a rate of 10 µL/min, delivering 100 µL of 0.1 mg/mL antibodies for 2 minutes to ensure appropriate preconcentration behavior. The selected flow channel was then prepared by flowing 1x PBS until a stable baseline was achieved. To functionalize the channel, a coupling solution containing 20 mg each of EDC (1-ethyl-3-(3-dimethylaminopropyl) carbodiimide hydrochloride, Life Technologies, Catalog no. 22980) and Sulfo-NHS (N-hydroxysulfosuccinimide, Life Technologies, Catalog no. 24510) in 700 µL of 100 mM MES at pH 4.7 was pushed through the chip for 15 minutes at the same flow rate. Following this, approximately 400 µL of the antibody solution (0.1 mg/mL in 10 mM MES pH 6.0) was applied at 10 µL/min for 25 minutes. The reaction was quenched by flowing 0.1 M ethanolamine (Life Technologies, Catalog no. 022793.30) for 2 minutes. The channel was then washed with PBS until a stable baseline was reestablished. The extent of antibody functionalization was assessed by measuring the baseline shift before and after conjugation, with an expected increase of over 2000 response units (RU) indicating successful antibody coupling.

Baseline establishment and experimental conditions for surface plasmon resonance (SPR) measurements were conducted as follows: Initially, the baseline was recorded to ensure stability, defined as a drift rate of less than 0.1 response units (RU) per minute. If the drift exceeded this threshold, the running buffer was flowed overnight or until the baseline stability criteria were met.

Upon achieving a stable baseline, experimental procedures commenced. Two flow rates, 1 μL/min and 10 μL/min, were chosen to cover a decade of flow rate variation while minimizing sample volume consumption. At the start of each cycle, the system was first set to a flow rate of 10 μL/min: the running buffer was flowed for five minutes, followed by a sample injection in high-performance mode for two minutes. Subsequently, the system was washed with the running buffer for 60 seconds before sequentially injecting Glycine-HCl (10 mM, pH 2) and 1% albumin, each for 20 seconds. The flow rate was then reduced to 1 μL/min, and the running buffer was flowed for an additional 30 minutes, followed by a five-minute sample flow. The flow rate was subsequently restored to 10 μL/min for the 2 final wash steps and injection sequences identical to the initial set. For both flow rates, the slope of the sensor response between 30-120 seconds post-injection was recorded to assess binding characteristics.

To correct for any system artifacts, a blank cycle was performed immediately following the sample measurements, using phosphate-buffered saline (PBS) instead of the sample to simulate identical injection conditions. The process was identical to that of the sample injections, including the adjustment of flow rates and the sequence of buffer and reagent injections. The measured slopes from the sample and blank cycles were compared to ascertain the specific binding response. These steps were repeated for each new sample, with the resultant signal representing the differential between the slopes obtained during sample and blank measurements, thus providing a corrected and reliable measure of the binding interactions. Based on the mass transfer constant in a laminar flow, the size of sEVs, the size of the sensor chip, and the diffusivity of sEVs, they are converted to sEV number concentrations.

### Cell media preparation

DiFi cells, derived from human colorectal carcinoma, were cultured in a three-dimensional (3D) system to replicate the in vivo tumor microenvironment closely. The 3D scaffolds, constructed from type-I collagen at a concentration of 2 mg/mL, were layered in a tripartite structure: basal and top layers of pure collagen flanked a central layer embedding DiFi cells at a density of 5,000 cells/mL. This configuration was incubated at 37°C in a humidified atmosphere of 5% CO_2_. Culture medium was supplemented with 10% fetal bovine serum (FBS), 2 μg/mL normocin, insulin-transferrin-selenium, epidermal growth factor, hydrocortisone, and T3 thyroid hormone, and refreshed every two to three days.

The human melanoma cell line A375P (RRID: CVCL_6233) and the human breast cancer cell line MDA-MB-468 (RRID: CVCL_0419) were maintained in high glucose Dulbecco’s Modified Eagle Medium (DMEM, Gibco, USA) enriched with 10% v/v EquaFetal Serum (Atlas Biologicals, USA), 2 mM L-glutamine, 100 U/mL penicillin-streptomycin, and 1 mM sodium pyruvate. These cells were cultured under standard conditions at 37°C in a 5% CO_2_ humidified atmosphere, ensuring optimal growth and maintenance.

Mouse fibroblast cells (3T3) were cultured in Minimum Essential Medium (MEM) supplemented with 10% FBS and 1% Antibiotic-Antimycotic Solution. The cells were housed at 37°C in a humidified 5% CO_2_ environment. Passaging involved washing with 1X phosphate-buffered saline (PBS), trypsinization with trypsin-EDTA, and a recovery period of at least one day before experimental use.

SH-SY5Y human neuroblastoma cells were cultured in Dulbecco’s Modified Eagle Medium/Nutrient Mixture F-12 (DMEM/F-12, Gibco) complete medium supplemented with 10% FBS and 1% Antibiotic-Antimycotic Solution at 37°C in a 5% CO_2_ humidified atmosphere.

GBM9 glioblastoma cells were cultured as neurospheres in Neurobasal medium devoid of serum (Gibco) and supplemented with 3 mM GlutaMAX, 1x B-27 supplement, 0.5x N-2 supplement, 20 ng/mL EGF (R&D Systems, MN), 20 ng/mL FGF (PEPROTECH, NJ), and 1% Antibiotic-Antimycotic Solution (Corning). Passaging was performed using the NeuroCult Chemical Dissociation Kit-Mouse (Stemcell Technologies, Canada) following the manufacturer’s guidelines.

## Supporting Information

Supporting Information is available from the Online Library or from the author.

## Acknowledgements

We want to thank Prof. Crislyn D’Souza-Schorey, Department of Biological Sciences, University of Notre Dame, for providing the melanoma cell line media samples. We would also like to thank the Notre Dame Biophysics Instrumentation Core Facility for the use of the Biacore T200 SPR system, which was purchased with funding from the NIH (S10 OD028553). This work was partially supported by the NIH Commons Fund, through the Office of Strategic Coordination/Office of NIH Director, 1UH3CA241684-01 (HCC & SS). R.J.C. acknowledges R35CA197570 and P50CA236733 from the National Cancer Institute, as well as the generous support from the Nicholas Tierney Memorial GI Cancer Fund.

## Author contributions

S.K., S.S., and H.C.C. conceived the idea; S.K., J.A.S., and T.S. acquired all data; S.K., S.S., H.C.C., J.L.F., and R.J.C. performed formal analysis; S.S. and H.C.C. acquired the funding and organized the project; S.K. and S.S. designed the experiments; S.K. conducted all IEMS experiments and characterization, including clinical samples in blind; S.K., G.G., and S.S. fabricated the sensor, microfluidic chip, and functionalized the sensor; S.K. and J.A.S. functionalized the SPR chip and performed the SPR experiments; S.K. and T.S. performed the ELISA experiments; G.K. and R.Z. provided the breast cancer, Neuroblastoma, and fibroblast cell lines; J.N.H., Q.Z., D.K.J., O. T., and M.X. provided the DiFi cell lines and purified DiFi EVs, supermeres, and exomeres samples; S.K. developed the figures and drafted the original manuscript. S.S., H.C.C., Y.Z., J.L.F., Q.Z., A.C., and R.J.C. reviewed and edited the manuscript with input from all authors.

## Data Availability Statement

The data that support the findings of this study are available from the corresponding author upon reasonable request.

## Supplementary Material for

### Estimating Voltage-Shift Signal Dependence on IEM Sensor Membrane Size

It is difficult to fabricate IEM sensors of the same size and hence the signal needs to be properly corrected for variations in the membrane size. In earlier reports (*44,45*), we and others have shown that, for an ideally selective membrane and a symmetric electrolyte with equal diffusivity and valency, the electromigration flux of the counterions is equal to its diffusive flux and hence the total ion flux is twice the diffusive flux. At the limiting current condition for a small membrane (or its nanoslot model), an ion-depleted region exists on the surface of the membrane with a dimension close to the size (radius) *R* of the membrane (*46*). The finite size of the depleted region is due to radial focusing of the diffusive flux to a point-like sink that is the membrane sensor. It is related to the fundamental solution 1/r of the diffusion equation in spherical coordinates. This known depletion or diffusion length allows us to estimate the limiting ion flux density as 2*Dc* /*R* and the Levich limiting current density *i*_lim_ as 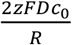. The limiting current is very easy to measure for each sensor, as it corresponds to a distinct voltage-independent current plateau of the I-V curve (see figure 2b in the text). The voltage shift is measured at the onset of the over-limiting current (*47*) from the limiting current where this relationship still holds (*48*). Since the over-limiting current, I-V curve is linear, the inverse scaling with respect to the membrane size *R* also holds for the voltage shift. In supplementary figure 1, we verify this *R*^-1^ relationship between the voltage shift observed and the size *R* of the membrane given as ∼ *i*_lim_/2*DzFc*_0_ where *i*_lim_ is the limiting current density, D is the diffusivity of the ions (1.5 × 10^−9^ *m*^2^/*s*), z is the valency of the ions (*z* = 1), *F* is the Faraday constant and *c*_0_ is the ionic concentration of the bulk (10 mM) during IV measurement.

**Figure S1.**
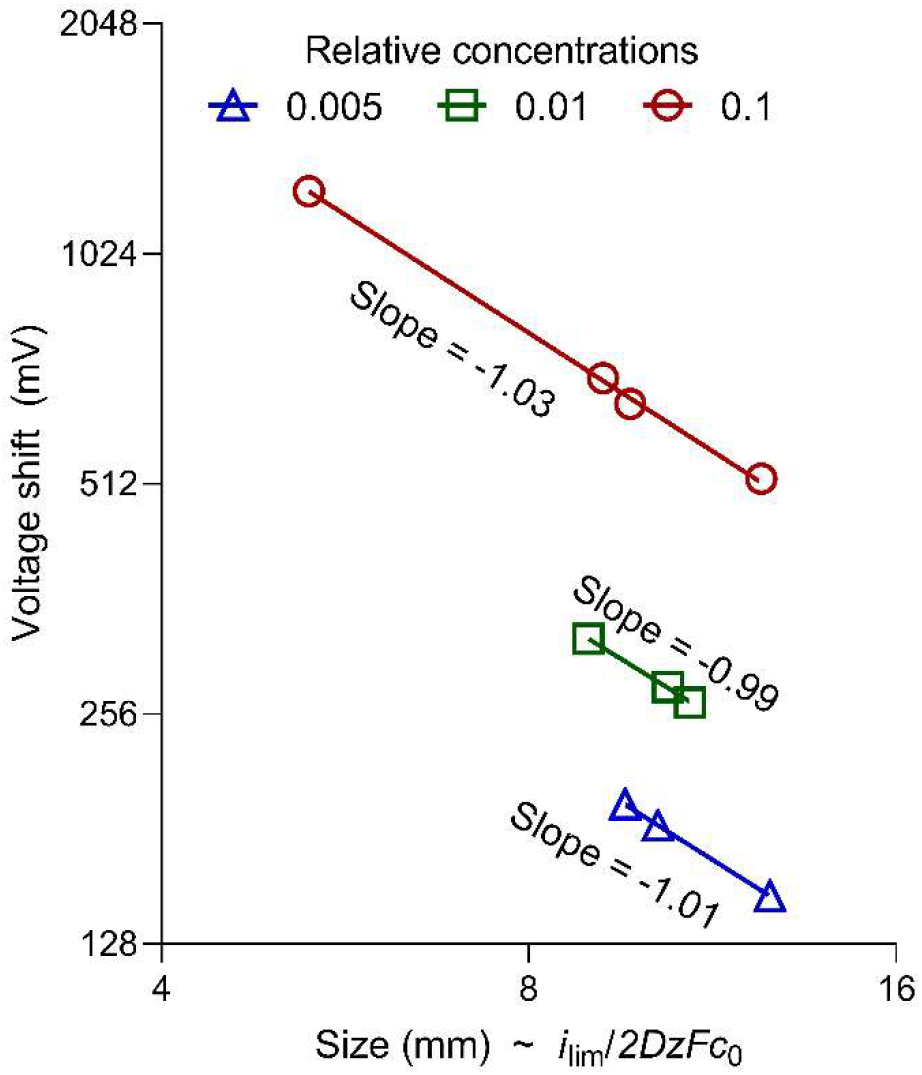
Size-dependence on the theoretical IEMS size of the voltage shift observed with respect to different stock concentrations of supermeres at different dilutions using anti-TGFBI antibody

The inverse linear relationship between the voltage shift and theoretical size of the IEMS sensor *R* suggests an opposing electric field generated by our analyte at a distance of about one membrane sensor distance, which is also the radius of our ion depletion zone. In this region, the ionic concentration reaches the lower asymptote of deionized water-like condition (10^−7^ M). Therefore, our signal is thus defined as representative of this charge given as (with initial state representing pre-incubation and final state representing incubation followed by wash), with *V*_1_ and *V*_2_ measured at the 2x limiting current for reference and to ensure we are in the over-limiting region. The parameter ϵ_0_ represents the permittivity of vacuum and ϵ_r_ the relative permittivity of water (∼80):

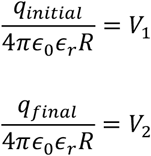

These equations again verify that the voltage shift scales inversely with *R*, with a proportionality constant that corresponds to the charge of the captured analyte. Therefore, the membrane size *R* dependence of the charge signal *S* can be replaced by the total limiting current 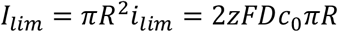:

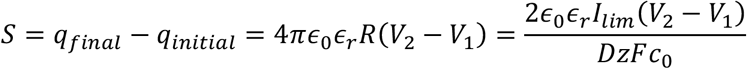

Here, *V*_2_ − *V*_1_ is essential for our voltage shift Δ*V*, so we can write it in final form as:

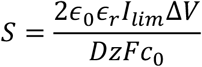

The value of *I*_lim_ is obtained from the I-V curve of every measurement.

